# Combining rare and common genetic variants improves population risk stratification for breast cancer

**DOI:** 10.1101/2023.05.17.23290132

**Authors:** Alexandre Bolze, Daniel Kiser, Kelly M. Schiabor Barrett, Gai Elhanan, Jamie M. Schnell Blitstein, Iva Neveux, Shaun Dabe, Harry Reed, Alexa Anderson, William J. Metcalf, Ekaterina Orlova, Ildiko Thibodeau, Natalie Telis, Ruomu Jiang, Nicole L. Washington, Matthew J. Ferber, Catherine Hajek, Elizabeth T. Cirulli, Joseph J. Grzymski

## Abstract

**Purpose:** In the United States, breast cancer clinical risk assessments are inconsistent and inequitable. The previous success of *BRCA1* and *BRCA2* screening has demonstrated that genetics could be used to reduce these inequities, if they are available and accessible across the population. The aim of this study is to evaluate the performance of different genetic screening approaches to identify women at high-risk of breast cancer in the general population.

**Methods:** We did a retrospective study on 25,591 women participating in the Healthy Nevada Project. Electronic Health Records (EHR) data were used to identify women with a family history of, and women who were diagnosed with breast cancer. The genetic analysis assessed the role of rare predicted loss-of-function (pLOF) variants in *BRCA1*, *BRCA2*, *PALB2*, *ATM* and *CHEK2*, as well as the combined role of common variants via a polygenic risk score. Women were considered at high-risk of breast cancer if they had greater than 20% probability of being diagnosed with breast cancer by age 70.

**Results:** Family history of breast cancer (FHx-BrCa) was ascertained on or after the record of breast cancer for 78% of women with both, indicating that this method for risk assessment is not being properly utilized for early screening. Genetics offered an alternative method for risk assessment. 11.4% of women in HNP were at high-risk of breast cancer based on their genetic risk: having a pLOF variant in *BRCA1*, *BRCA2* or *PALB2* (hazard ratio = 10.4, 95% confidence interval: 8.1-13.5), or a pLOF variant in *ATM* or *CHEK2* (HR = 3.4, CI: 2.4-4.8), or being in the top 10% of the polygenic risk score distribution (HR = 2.4, CI: 2.0-2.8). We also showed that combining PRS with pLOF in *ATM* and *CHEK2* allowed a better identification of participants with high risk while minimizing false positives. Women with a pLOF in *ATM* or *CHEK2* and in the top 50% of the PRS are at high risk (39.2% probability of breast cancer at age 70), while those with a pLOF in *ATM* or *CHEK2* and in the bottom 50% of the PRS are not at high risk (14.4% probability of breast cancer at age 70).

**Conclusion:** These results suggest that a combined monogenic and polygenic approach may best capture the inherited risk for breast cancer across the population.

## Introduction

Disparities pervade all aspects of breast cancer screening and treatment. Current clinical risk assessments, aimed at identifying women eligible for referral to high-risk breast cancer clinics, are often inconsistently applied, leading to inequitable outcomes ^1–3^. In 2021, Nevada attempted to address this issue by enacting a law, SB251, mandating payors to cover counseling and genetic testing for women identified as high-risk for breast cancer. However, consistent implementation of screening and precise capture of family history data by all providers remains a crucial first step in assessing who should receive counseling and genetic testing.

An alternative approach to identifying individuals who may benefit from referral to a high-risk clinic is to adopt a population-level genetic screening approach, sequencing all women ^4–7^. The detection of rare, high-impact variants, such as loss of function variants in *BRCA1* and *BRCA2*, is used to identify individuals who should undergo mammography screening at an earlier age and with greater frequency ^4, 8^, and consider risk-reducing measures. Variants in other genes, including *PALB2*, *ATM* and *CHEK2*, are also strongly associated with breast cancer risk ^9–12^. However, a significant association with breast cancer may not warrant screening the entire population, especially if the positive predictive value in the general population is low^13^. Additionally, polygenic risk scores (PRS), based on common variants associated with breast cancer, have been developed and replicated across numerous cohorts ^14, 15^. In the United Kingdom, a score based on 313 SNPs has been integrated in BOADICEA to create a comprehensive breast cancer risk prediction model ^16^, which aims to identify individuals with high or low risk for breast cancer. Despite the apparent benefits of using genetic information to identify women at high risk of breast cancer, particularly women with incomplete family history, many questions remain about the real-world impact of such a measure and the most equitable way to implement it across the population in the United States.

The Healthy Nevada Project (HNP) is an all-comer genetic screening and research project based in northern Nevada ^4^. HNP participants with CDC Tier 1 findings, including hereditary breast and ovarian cancer (HBOC), are notified and provided with genetic counseling. All participants consent to a research protocol that encompasses: (i) clinical Exome+® sequencing, (ii) linking of sequencing data with available electronic health records, (iii) return of secondary findings, and (iv) recontact for health surveys and participation in future studies. This study offers opportunities to retrospectively evaluate various genomic screening methods for identifying women at high risk of breast cancer, and to prospectively assess the impact and outcomes of these methods when applied in a clinical setting.

The objectives of this study are to: (i) Evaluate the effectiveness of family history as a tool for identifying women at high risk of breast cancer. (ii) Determine the impact of pathogenic variants in *BRCA1*, *BRCA2*, *PALB2*, *ATM*, and *CHEK2*, as well as the presence of a high polygenic risk, on breast cancer diagnosis within the HNP cohort. (iii) Characterize the population of women newly identified or missed based on the utilization of different combinations of genetic and family history information. Many studies and institutions have their own definitions and thresholds for what qualifies as ‘high risk’ for breast cancer^16–18^. In this study, we define ‘high risk’ of breast cancer as a 20% or greater risk of breast cancer by age 70, corresponding to >2X the average risk for women in the United States.

## Results

### Demographics of the Healthy Nevada Project

As of December 2022, 47,179 individuals had consented to and been sequenced as part of the Healthy Nevada Project^4, 13^. Our analysis focused on women with available electronic health records (EHR) from Renown Health. In total, we analyzed the genetic and clinical information of 25,591 participants (**Table 1**). The mean age was 53.8 years, with a bimodal age distribution (**Figure S1A**). Of these participants, 4,977 (19.4%) were 70 years or older in 2022. We utilized EHR diagnosis tables to identify participants diagnosed with breast cancer. In total, 1,295 women (5.1%) had at least one ICD10-CM code starting with C50 (indicating malignant neoplasm of the breast), D05 (indicating carcinoma in situ of the breast) or Z85.3 (indicating personal history of malignant neoplasm of the breast) (**Table 1**, **Figure S1B-C**). Among those 70 years of age or older in 2022, 570 (11.5%) had a diagnosis of breast cancer. This prevalence is consistent with estimates from the National Cancer Institute, reporting that 12.9% of women born in the United States today will develop breast cancer at some time during their lives ^19^ .

**Table 1:**
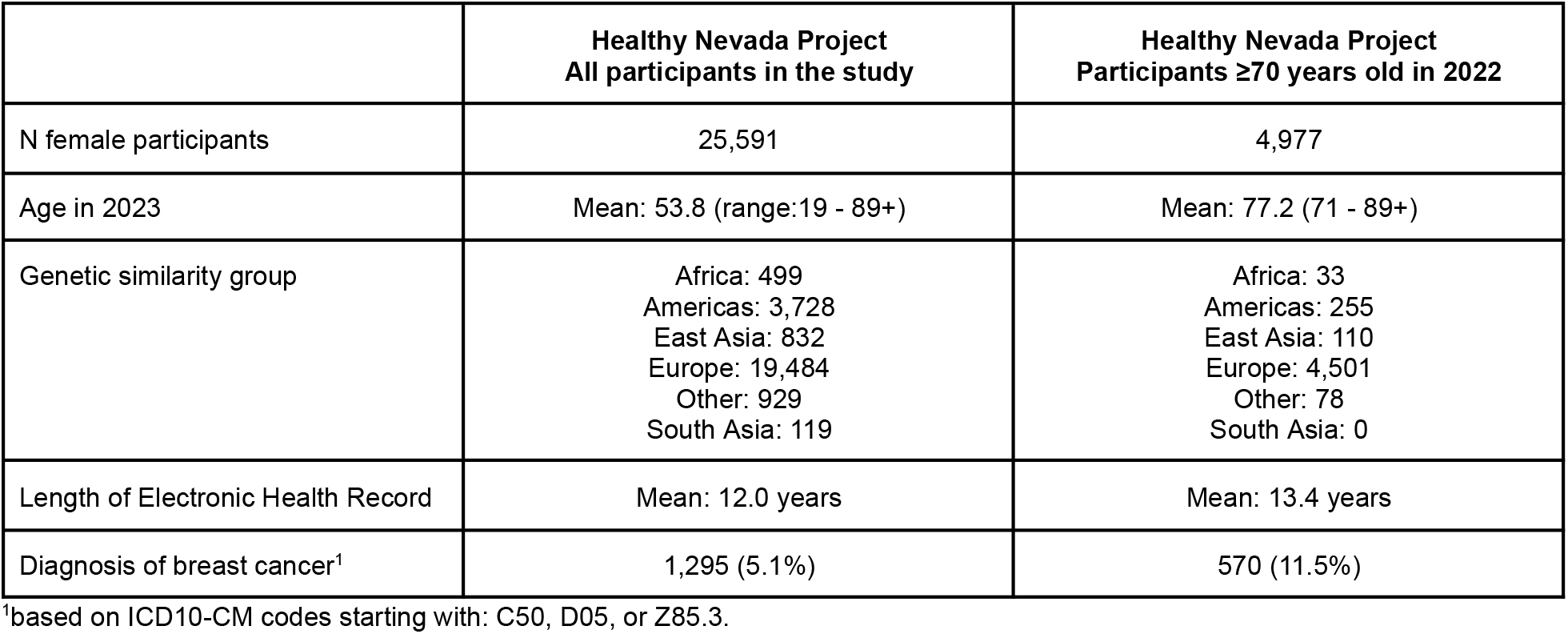
Demographics of participants from the Healthy Nevada Project included in this study.

### Family history misses most women at increased risk of breast cancer prior to diagnosis

Current clinical guidelines heavily depend on family history to identify women at increased risk for breast cancer ^20^. We identified three sources of breast cancer family history records in the EHR: diagnosis codes, a family history table, and a table containing responses to the seven-question family history screening that was implemented in 2021 to comply with SB251. We evaluated the agreement between these three sources for identifying patients with a family history of breast cancer (FHx-BrCa). Diagnosis codes captured 97.6% of all participants with any positive FHx-BrCa in any of these three tables (**Figure S2**). Therefore, we used only diagnosis codes to identify women with a FHx-BrCa in subsequent analyses. A total of 1,772 women (6.9%) had at least one ICD10-CM Z80.3 code indicating FHx-BrCa. The mean age of the first code was 50.2 years (**Figure 1A**). However, the mean age of the first FHx-BrCa code for women diagnosed with breast cancer was 58.6 years, closer to the mean age for breast cancer diagnosis (60.2 years) (**Figure 1A**), suggesting family history may be taken at the time of diagnosis.

We next examined the specific timing of the family history record and breast cancer diagnosis (**Figure 1B**). For the majority of participants with both a family history and a diagnosis of breast cancer, the family history ascertainment was recorded at the same time or after the breast cancer diagnosis (**Figure 1B**). There were 30% of first FHx-BrCa records simultaneous with the first breast cancer diagnosis record (between 1 week before and 1 month after) and 48% occuring afterward (1 month to 15 years after) (**Figure 1B**). As a control, we looked at the timing of the record of FHx-BrCa and of a diagnosis of another common unrelated disease. There was no enrichment for the family history code to be at the same time as the diagnosis of T2D (Chi-square test for comparison to enrichment seen between FHx-BrCa and BrCa diagnosis, p= 1.9e-11), or other common non-cancer conditions with a similar average age of onset to breast cancer (**Figure 1C**). This suggests that the bias observed around breast cancer diagnosis was not due to a general effect of receiving more information about a patient at certain time points, such as the first encounter at the health system. We next looked at codes for family history of other conditions. 26.8% of women had at least one FHx code not for FHx-BrCa code. Among those with FHx-BrCa recorded simultaneously or after the first BrCa diagnosis, 14% (35 of 259) had at least one FHx code for a condition other than BrCa recorded before the BrCa diagnosis (**Figure 1D**, **Figure S2C**), indicating the bias observed was likely due to an ascertainment bias rather than a bias in recording information in the EHR. These results demonstrate that a patient is more likely to be ascertained for FHx-BrCa on or after a diagnosis of breast cancer or suspicion of breast cancer.

**Figure 1:**
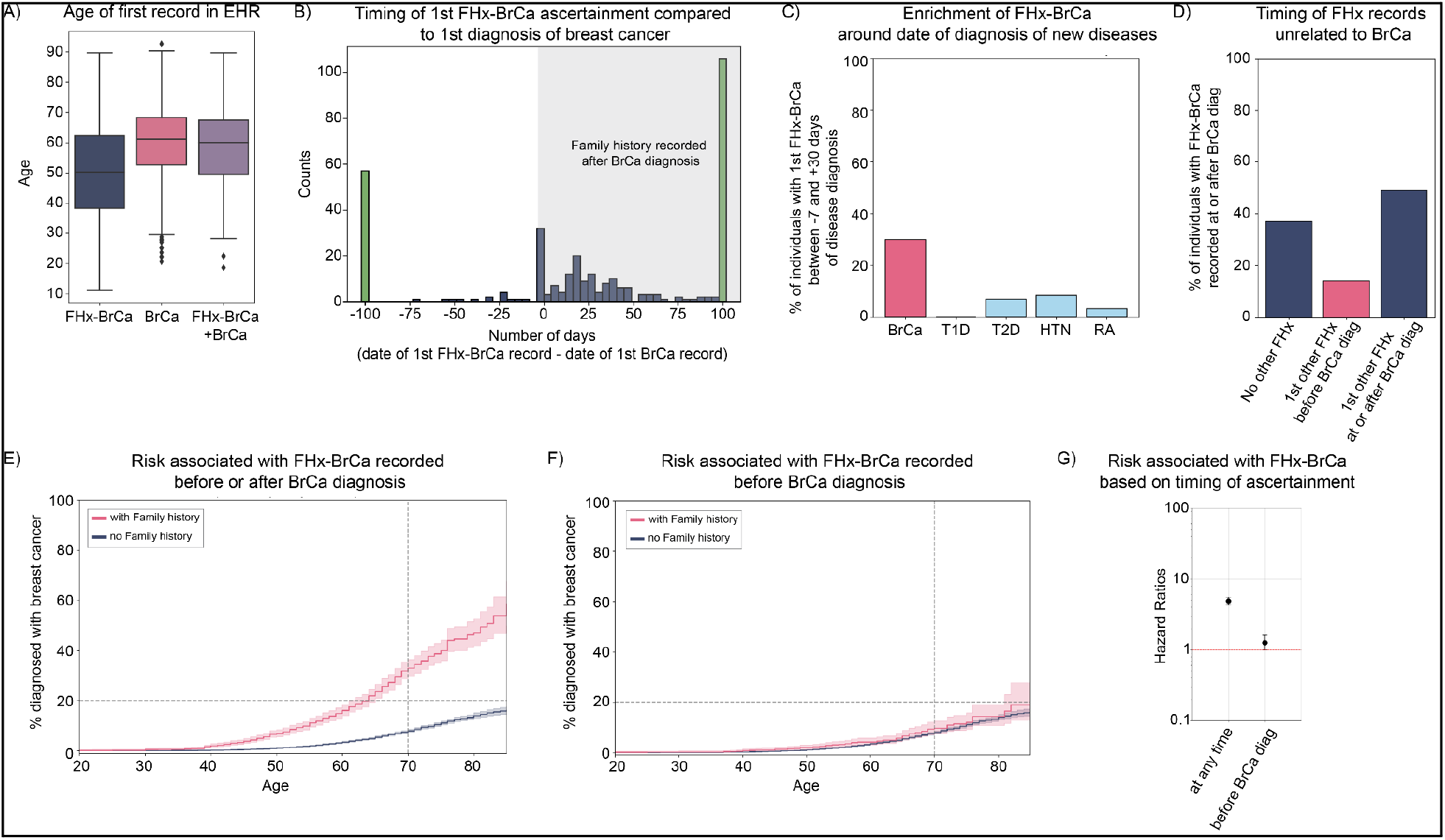
Impact of family history of breast cancer (FHx-BrCa) on risk of breast cancer (BrCa). **(A)** Mean age of first record in the electronic health records. N= 1,772 for FHx-BrCa; N= 1,295 for BrCa; N= 331 for FHx-BrCa and BrCa. **(B)** Days between 1st record of FHx-BrCa and 1st record of BrCa. All bars on the left side (negative numbers) indicate individuals for whom FHx-BrCa was recorded before a BrCa diagnosis. Green bars indicate points before -100 days, or after +100 days. **(C)** Percentage of individuals with the 1st record of FHx-BrCa within a time range of -7 to +30 days around the 1st diagnosis of BrCa (pink bar) or other conditions (light blue bars). From left to right: type 1 diabetes (T1D), type 2 diabetes (T2D), hypertension (HTN) and rheumatoid arthritis (RA). **(D)** Timing of 1st family history (FHx) record other than FHx-BrCa for individuals with the 1st FHx-BrCa recorded within 1 week or after the 1st BrCa record. N=259. **(E)** Kaplan Meier curves showing the % of women with a breast cancer diagnosis by age. Pink curve: with FHx-BrCa, N= 1,772. Blue curve: without FHx-BrCa, N=23,819. **(F)** Kaplan Meier curves showing the % of women with a breast cancer diagnosis by age. Participants with a 1st FHx-BrCa recorded within 1 week of 1st BrCa diagnosis or after 1st BrCa diagnosis were excluded from analysis. Pink curve: with FHx-BrCa, N= 1,513. Blue curve: without FHx-BrCa, N=23,819. (G) Cox proportional hazard ratios and their 95% Confidence Intervals. Y-axis is on a log scale.

Despite this limitation, we assessed the risk of developing breast cancer for women who were aware of and reported a family history of breast cancer. Among women with a FHx-BrCa code in their medical records, 18.7% (331 of 1,772) had a diagnosis of breast cancer, compared to 4.0% (964 of 23,819) of women without, and the hazard ratio (HR) was 4.9 (95% Confidence Interval (CI): 4.3-5.5) (**Figure 1E**). To minimize the impact of family history ascertainment bias, we next analyzed the risk associated with a positive family history of breast cancer after removing participants whose first FHx-BrCa code appeared simultaneously with or after the first diagnosis of breast cancer. In this scenario, 4.8% (72 of 1,513) of women with a positive family history had a diagnosis of breast cancer, a risk similar to women without a family history (HR=1.3, CI: 1.0-1.6, p=0.05, log-rank test) (**Figure 1F**). However, this analysis likely overcorrected the ascertainment bias, as it removed the majority of cases among the group with a family history and did not remove cases in the group without a family history. Overall, these results showed that at least 6.9% of women had a positive family history of breast cancer, but family history is often ascertained at or after the time of diagnosis. This (i) undermines the goal of identifying patients before diagnosis, and (ii) makes it challenging to measure the actual risk of developing breast cancer for those with a family history of breast cancer (**Figure 1G**).

### Loss of function variants in *PALB2*, *ATM* and *CHEK2* significantly increase risk of breast cancer

As an alternative to family history, we examined five genes – *BRCA1*, *BRCA2*, *PALB2*, *ATM* and *CHEK2* – where haploinsufficiency is known to be associated with breast cancer risk based on previous studies ^13, 16, 21, 22^. We thus considered high-confidence predicted loss-of-function pathogenic variants in these five genes and pathogenic variants in ClinVar reviewed by an expert panel. We also examined pathogenic deletions (copy number of 1 or 0) of one or more protein-coding exons of the MANE transcript in these genes. Details of variant annotation and classification are in the methods (**Figure S3A**). The variant calls for these genes were of high quality (**Figure S3B-C**), and all variants annotated as pathogenic had an allele frequency below 0.1% in the population, with the exception of the well-known *CHEK2* del1100C variant (**Figure S3D**). The list of identified pathogenic single-nucleotide variants or small indels is in **Table S1**, and the list of larger deletions is in **Table S2**. In total, we identified 410 (1.6%) women who had a heterozygous pathogenic variant in one of these five genes (**Table S3**). No individual had a homozygous pathogenic variant or a copy number of zero in these genes. One individual had two heterozygous pathogenic variants in different genes (one in *BRCA1* and one in *BRCA2*).

The risk of breast cancer was highest for women with a pathogenic variant in *BRCA1* or *BRCA2*: hazard ratio (HR) of 16.2 (95% CI: 10.9-24.1, p=2.6e-76) for *BRCA1* and HR = 8.5 (CI: 5.8-12.5, p=1.3e-39) for *BRCA2* (**Figure 2A**). The next-highest risk was for women having a pathogenic variant in *PALB2*, with a hazard ratio of 6.3 (95% CI: 3.4-11.7, p=3.2e-11), followed by *ATM*, with a hazard ratio of 4.3 (95% CI: 2.6-7.2, p=1.0e-09), and finally *CHEK2,* with a hazard ratio of 2.6 (95% CI: 1.7-4.2, p=2.4e-05) (**Figure 2A**, **Table S4**). The probability of diagnosis at age 70 was 9.3% for women without a pathogenic variant, 76% for women with a *BRCA1* pathogenic variant, 55% for those with a *BRCA2* pathogenic variant, 36% for those with a *PALB2* pathogenic variant, 37% for those with an *ATM* pathogenic variant and 19% for those with a *CHEK2* pathogenic variant. This analysis revealed that some individuals with a pathogenic variant in *PALB2* were diagnosed with breast cancer well before the age of 50 (**Table S4**) and had a similar risk to those with a *BRCA2* variant before the age of 50 (**Figure 2A**), when until recently, guidelines recommended beginning screening for everyone.

**Figure 2:**
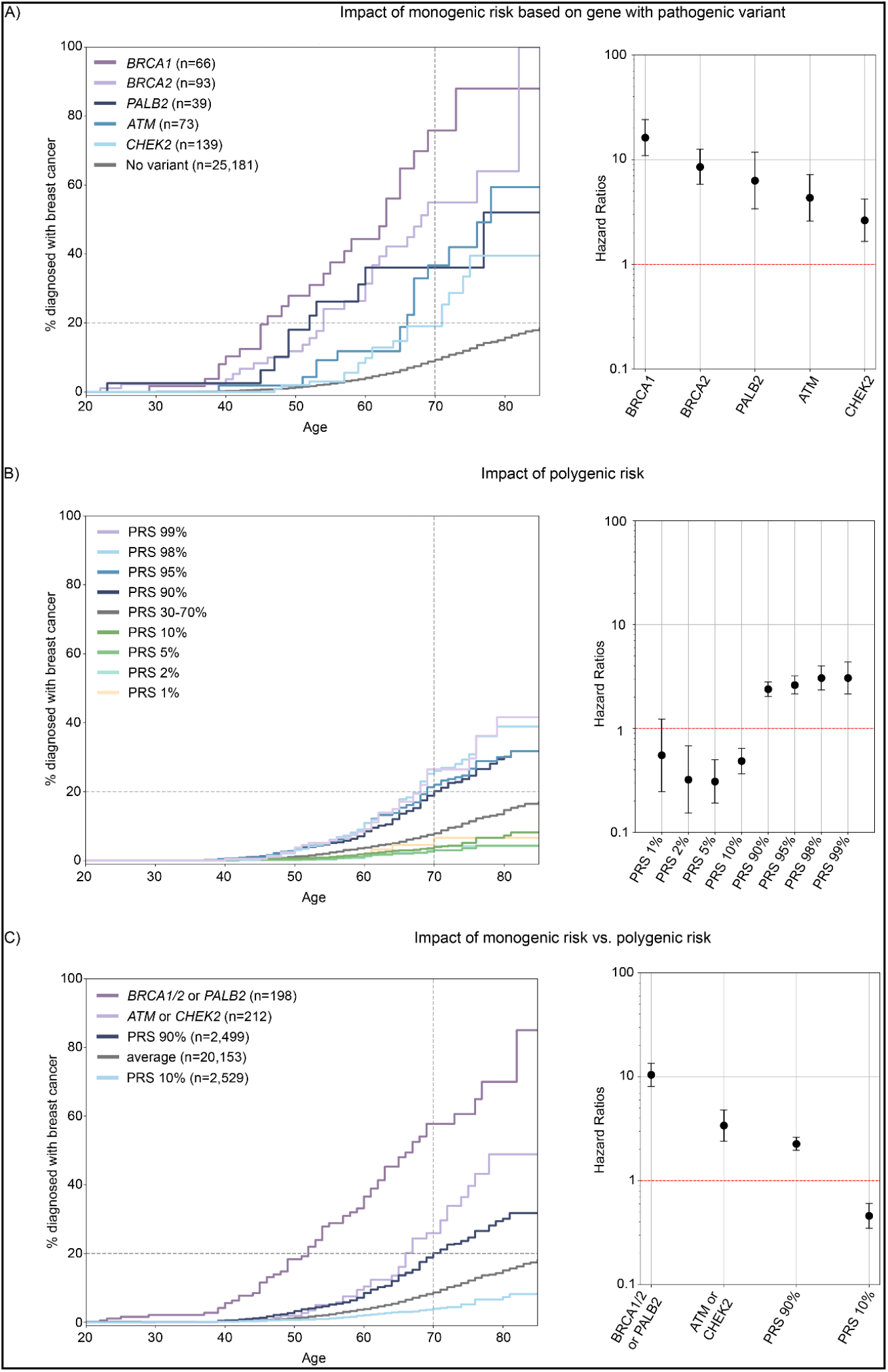
Impact of monogenic or polygenic risk on breast cancer diagnosis. **(A)** Left panel: Kaplan Meier curves showing the % of women with a breast cancer diagnosis by age based on whether they have a pathogenic variant in one of 5 genes: *BRCA1* (purple curve), *BRCA2* (light purple), *PALB2* (dark blue), *ATM* (blue), or *CHEK2* (light blue). Gray curve: no pathogenic variants. Right panel: Cox proportional hazard ratios and their 95% Confidence Intervals; Y-axis is on a log scale. **(B)** Left panel: Kaplan Meier curves showing the % of women with a breast cancer diagnosis by age based their breast cancer polygenic risk score: PRS 99% (top 1%) in light purple, PRS 98% (top 2%) in light blue, PRS 95% (top 5%) in blue, PRS 90% (top 10%) in dark blue, PRS 30-70% (average score) in gray, PRS 10% (bottom 10%) in dark green, PRS 5% (bottom 5%) in green, PRS 2% (bottom 2%) in light green, and PRS 1% (bottom 1%) in yellow. Right panel: Cox proportional hazard ratios and their 95% Confidence Intervals; Y-axis is on a log scale. **(C)** Left panel: Kaplan Meier curves showing the % of women with a breast cancer diagnosis by age based on their monogenic or polygenic risk. Purple curve: women with a pathogenic variant in BRCA1 or BRCA2 or PALB2. Light purple curve: women with a pathogenic variant in ATM or CHEK2. Dark blue curve: women without a pathogenic variant who are in the top 10% of the PRS distribution. Light blue curve: women without a pathogenic variant who are in the bottom 10% of the PRS distribution. Gray curve: all other women. Right panel: Cox proportional hazard ratios and their 95% Confidence Intervals; Y-axis is on a log scale.

### High polygenic risk significantly increases risk of breast cancer, but less so than monogenic risk

Next, we evaluated the clinical impact of having a high polygenic risk for breast cancer, which quantifies the contribution of common variants known to be associated with this disease. We used a validated polygenic risk score (PRS) of 313 SNPs (**Table S5**) ^14^. Quality control measures for the SNPs used are depicted in **Figure S4 A-B**, and implementation details can be found in the Methods section. The median breast cancer PRS showed variation based on genetic similarity (**Table S6**). When comparing breast cancer cases and controls, the distributions of the PRS were shifted (**Figure S4 C-D**), and the PRS achieved an AUC of 0.63 in participants in the Europe genetic similarity group and 0.66 in participants in the Americas genetic similarity group, consistent with findings reported in other studies using this PRS^14^. While we could not compute the AUC for other genetic similarities due to an insufficient number of participants, subsequent analyses included all participants regardless of their genetic similarity. To enable the study of all participants, and, to better emulate a real-world scenario where all participants would receive a result, we assigned a PRS percentile to each participant based on their rank within their genetic similarity group. We then combined all the participants and assessed the impact of being in the top or bottom of their group-specific PRS distribution. Furthermore, we excluded all participants having a pathogenic variant in one of the 5 genes previously studied, namely *BRCA1*, *BRCA2*, *PALB2*, *ATM* and *CHEK2*, to ensure that these variants would not confound the results.

Women in the top 2% and top 10% of their group-specific PRS distribution exhibited an increased risk of breast cancer compared to women with an average polygenic risk, with hazard ratio of 3.1 and 2.4 respectively (CI: 2.3-4.0 and 2.0-2.8, p=6.4e-18 and 1.9e-28 respectively) (**Figure 2B**, **Table S7-S8**). Both of these risks were lower compared to the risk associated with having a pathogenic variant in *BRCA1*, *BRCA2* or *PALB2* (HR=10.4, CI: 8.1-13.5), but were closer to the risk associated with having a pathogenic variant in *ATM* or *CHEK2* (HR=3.4, CI: 2.4-4.8) (**Figure 2C**). For women with a PRS in the top 2% and top 10% of their group-specific distribution and without a pathogenic variant, the probability of diagnosis at age 70 was 26% and 20% respectively. Interestingly, the probability of diagnosis at age 70 was just 3.9% for women with a PRS in the bottom 10% of the group-specific distribution and without a pathogenic variant (**Table S8**).

### Polygenic risk can modify monogenic risk for genes of intermediate penetrance: *ATM* and *CHEK2*

Next, we investigated whether we could enhance the predictive power by combining polygenic risk with other risk factors. To improve statistical power and avoid overly small subgroups, we partitioned the cohort into two groups based on PRS. The hazard ratio of women in the top 50% of the PRS compared to those in the bottom 50% of the PRS was 2.1 (CI: 1.8-2.3) (**Figure 3A, Table S9**). While most combinations of polygenic risk with monogenic risk (**Figure 3B-C**), polygenic risk with family history (**Figure 3D**) or family history with monogenic risk (**Figure 3E-F**) enhanced risk stratification, only one combination – PRS and pLOF in *ATM* or *CHEK2* – appeared clinically useful. This was because individuals in the higher risk group were clearly above the 20% risk at age 70, a threshold that defines women at high-risk^17^, while the lower risk group was clearly below this threshold. The probability of breast cancer diagnosis at age 70 in individuals with a pathogenic variant in *ATM* of *CHEK2* and in the top 50% of the PRS distribution was 39.2%, while those in the bottom 50% of the PRS distribution had a risk of only 14.4% (**Figure 3C**, **Table S9**). Individuals with a pathogenic variant in *BRCA1*, *BRCA2* or *PALB2* had a risk of breast cancer above 20% at age 70, regardless of their PRS (**Figure 3B**) or family history (**Figure 3E**, **Table S9**).

Previous studies have demonstrated a correlation between family history and PRS, prompting a reevaluation and adjustment of some weights in the PRS to account for this relationship ^14, 23^. Consequently, we sought to investigate if there were associations between PRS, rare pathogenic variants, and family history in our data. To probe these potential correlations, we excluded individuals with a breast cancer diagnosis and limited the analysis to the group with European genetic similarity to utilize normalized PRS values (instead of percentiles). We discovered a significant association between having a family history of breast cancer and possessing a pathogenic variant (p=3.5e-9, Fisher’s exact test and OR=3.5). Additionally, we found a positive and significant association between normalized PRS values and having a family history of breast cancer (p=4.6e-09, t-test), but no association between normalized PRS values and having a pathogenic variant in any of the five genes associated with monogenic risk (p=0.74, t-test). Collectively, these findings suggest that incorporating both PRS and monogenic risk in genes of intermediate penetrance may be advantageous. Conversely, integrating a family history of breast cancer with other risk factors may yield reduced benefits, partially due to the connection between family history of breast cancer with both monogenic risk and PRS.

**Figure 3:**
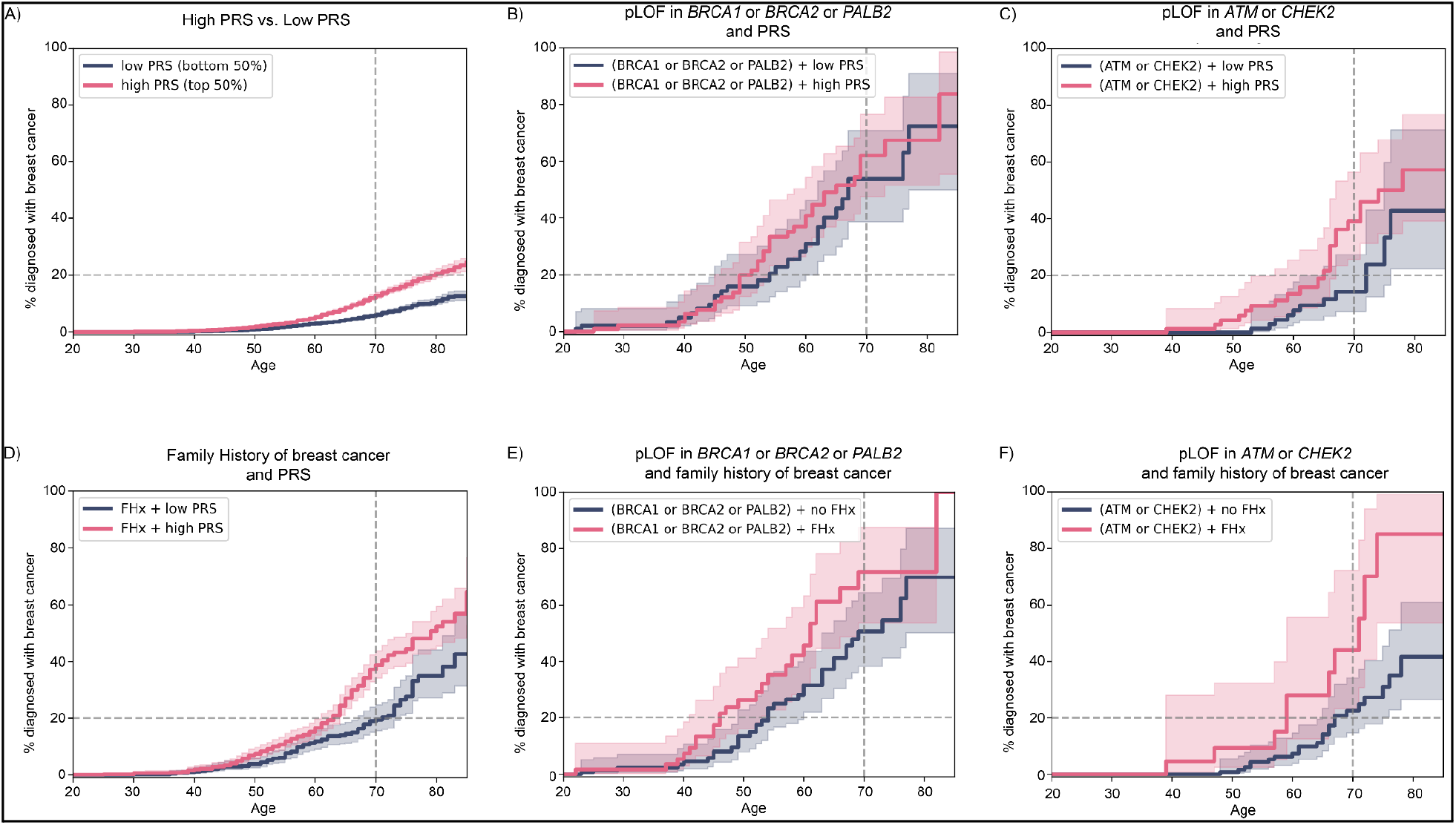
Impact of combining two non-modifiable risk factors on breast cancer diagnosis. Kaplan Meier curves showing the % of women with a breast cancer diagnosis by age based on **(A)** being in the top 50% of the PRS (N=12,594; pink curve) vs. bottom 50% of the PRS (N=12,587; blue curve). **(B)** having a pathogenic variant in *BRCA1*, *BRCA2* or *PALB2* and a high PRS (N=100; pink) vs. having a pathogenic variant in *BRCA1*, *BRCA2* or *PALB2* and a low PRS (N=98; blue). **(C)** having a pathogenic variant in *ATM* or *CHEK2* and a high PRS (N= 112; pink) vs. having a pathogenic variant in *ATM* or *CHEK2* and a low PRS (N= 100; blue). **(D)** having a family history of breast cancer and a high PRS (N=986; pink) vs. having a family history of breast cancer and a low PRS (N=700; blue). **(E)** having a pathogenic variant in *BRCA1*, *BRCA2* or *PALB2* and a family history of breast cancer (N=61; pink) vs. having a pathogenic variant in *BRCA1*, *BRCA2* or *PALB2* and no family history of breast cancer (N=137; blue). **(F)** having a pathogenic variant in *ATM* or *CHEK2* and a family history of breast cancer (N=25; pink) vs. having a pathogenic variant in *ATM* or *CHEK2* and no family history of breast cancer (N=187; blue). 95% Confidence Intervals are represented in light pink or blue.

### Evaluation of different screening strategies based on the combination of family history risk, monogenic risk and polygenic risk

While determining the odds ratio for developing breast cancer based on screening results is crucial, implementing genetic screening in the general population necessitates considering additional factors. These include the number of individuals testing positive who will require further counseling and preventive screening, the number of individuals overlooked by current guidelines, and those diagnosed with breast cancer before the age of 50 who are missed by genetic screening strategies. Accordingly, we calculated these parameters for various genetic screening strategies and panels within the Healthy Nevada Project (**Table 2**). Based on these figures and current medical practices in the United States, one potential strategy could involve referring all women with either a family history of breast cancer, a pathogenic variant in *BRCA1*, *BRCA2* and *PALB2*, or a pathogenic variant in *ATM* and *CHEK2* coupled with a high PRS to a high-risk breast cancer clinic (**Figure S5A**). An alternative approach could entail discontinuing the ascertainment of family history of breast cancer and instead refer all women with a pathogenic variant in *BRCA1*, *BRCA2*, *PALB2*, *ATM* and *CHEK2* or a PRS in the top 10% (**Figure S5B**). It is noteworthy that a considerable number of patients diagnosed with breast cancer before the age of 50 would not be identified by any of these screening strategies. Incorporating other independent risk factors such as prior health history, dense breast tissue, and menstrual and reproductive history may aid in identifying some of these patients earlier^16, 24, 25^.

**Table 2:** Impact of different strategies for genetic screening in the entire population.

| Genetic strategy<br>genetic component<br>or family history | N<br>(% of<br>population) | Probability of being<br>diagnosed with breast<br>cancer at age 70<sup>1</sup> | N with early onset breast<br>cancer undetected by strategy<br>(% of those with a BrCa<br>diagnosis ≤50 yo)<sup>2</sup> | N with a family history<br>recorded at any time<br>(% of those identified<br>by genetic strategy) |
| --- | --- | --- | --- | --- |
| *BRCA1* + *BRCA2* | 159<br>(0.6%) | 63.1% | 209<br>(91.3%) | 53<br>(33.3%) |
| *BRCA1* + *BRCA2* + *PALB2* +<br>(*ATM* + *CHEK2*) * top 50% PRS | 298<br>(1.2%) | 51.1% | 201<br>(87.8%) | 72<br>(24.2%) |
| *BRCA1* + *BRCA2* + *PALB2* +<br>*ATM* + *CHEK2* | 410<br>(1.6%) | 40.4% | 201<br>(87.8%) | 86<br>(21.0%) |
| *BRCA1* + *BRCA2* + *PALB2* +<br>*ATM* + *CHEK2* + 10% PRS | 2,909<br>(11.4%) | 23.2% | 156<br>(68.1%) | 297<br>(10.2%) |
| No genetic test. Family History<br>best case scenario<sup>3</sup> | 1,772<br>(6.9%) | 32.9% | 139<br>(60.7%) | NA |
| No genetic test. Family History<br>worse case scenario<sup>4</sup> | 1,513<br>(5.9%) | 9.6% | 207<br>(90.4%) | NA |

<sup>1</sup> based on Kaplan Meier curves at age 70.
<sup>2</sup> There are 229 female participants (out of 25,591) who were diagnosed with breast cancer before the age of 50 in the study population.
<sup>3</sup> If all women with a FHx-BrCa recorded at any time were ascertained prior to diagnosis.
<sup>4</sup> Only including women with FHx-BrCa recorded in EHR before a BrCa diagnosis.

## Discussion

The recently updated guidelines from the US Preventive Services Task Force (USPSTF) now recommend biennial screening mammography for all women aged 40 to 74 (previously aged 50-74) at average risk. Guidelines suggest that women with a parent, sibling, or child diagnosed with breast cancer, or who have early onset breast cancer themselves, may benefit from earlier screening. Despite operational and practical challenges, the ascertainment of family history is recommended ^26, 27^. Challenges to ascertaining family history include time constraints during medical visits, smaller family sizes, families with adopted members, families that do not discuss health issues, and male-dominant pedigrees. In a large health system from Northern Nevada, we found a 6.9% prevalence of breast cancer family history. Other studies focusing on family history among women in the general population reported higher prevalence: 16.1% among 222,019 women in the Breast Cancer Surveillance Consortium^28^, and 24.1% based on the National Health Interview Survey^29^. We also noted an ascertainment bias, as family history of breast cancer often is determined at the time of breast cancer diagnosis. The true risk associated with having a family history of breast cancer is likely lower than the hazard ratio of 4.9 we found, once a larger number of individuals with a family history and less biased ascertainment are taken into account. Furthermore, the mean age at the first code for family history of breast cancer was 50.2 years, suggesting a missed opportunity for early-onset breast cancer screening^30^. Although family history ascertainment can be enhanced with tools such as the FHS-7, which is limited to seven questions, these tools often have low specificity and might provide an incomplete picture of cancer risk based on family history ^31, 32^. Our findings suggest that risk based on family history is ascertained too late and too infrequently.

Population genetic screening presents a promising alternative^4, 33^. Our results confirmed that rare pLOF variants in *BRCA1*, *BRCA2*, *PALB2*, *ATM* and *CHEK2* confer significant risk to breast cancer in the overall population ^7, 10, 12, 34, 35^. These results provide useful guidance for discussions on breast cancer risk for women with a variant in *ATM* or *CHEK2*, which are less studied. We found that by age 70, women with an *ATM* pLOF variant had a 37% accumulated risk of being diagnosed with breast cancer (HR = 4.3), aligning with the 28-38% lifetime risk mentioned in the NCCN guidelines^18^. Women with a *CHEK2* pLOF had a 19% accumulated risk (HR = 2.6), which is on the lower end of the 23-48% lifetime risk mentioned in the NCCN guidelines^18^. For *ATM* and *CHEK2*, the NCCN guidelines suggest an annual mammogram starting at age 40 but specify insufficient evidence to support risk-reducing mastectomy based on *ATM* or *CHEK2* variant status alone; instead, management should be based on personal risk factors and family history^18^. Our findings, however, show a significant correlation between family history and having a pathogenic variant, as well as between family history and a high polygenic risk score. Adding family history to genetic findings might not substantially increase predictive utility. However, our ability to evaluate this was hampered by the lack of an unbiased baseline family history ascertainment (e.g., a large group of women with family history data systematically collected at age 18). On the other hand, we showed that coupling the identification of a variant in *ATM* or *CHEK2* with the calculation of a polygenic risk score might effectively identify women most at risk of breast cancer.

To explore polygenic risk in the population, we employed a published and validated 313-SNPs PRS for breast cancer^14^ and confirmed this PRS model as a robust candidate for clinical implementation globally^36^. Individuals within the top decile of polygenic risk exhibit a 20.1% accumulated risk of breast cancer by age 70, corresponding to an approximate 2.5-fold increase in lifetime breast cancer risk. Despite this evidence supporting the value of a polygenic risk score as a screening tool, many studies have highlighted challenges associated with their clinical use ^37, 38^. These include: (i) the current absence of clinical trials to measure their efficacy and determine the most effective methods for communicating and returning polygenic risk results, (ii) the fact that most PRS have been built using data from individuals of Europe genetic similarity and may be less predictive for individuals from other backgrounds, and (iii) the larger effect of monogenic risk compared to polygenic risk. Screening for both rare pathogenic variants and polygenic risk, and combining these results, could address some of these challenges, a use case that has been discussed in other studies and perspectives ^38–40^. Based on our data from the Healthy Nevada Project, a possible screening strategy could focus on women with a pathogenic variant in *BRCA1* or *BRCA2* or *PALB2*, and then consider women with a pathogenic variant in genes with slightly lower penetrance (such as *ATM* or *CHEK2*) who also fall within the top 50% of the PRS distribution. This approach identifies women at much higher risk compared to current guidelines and can be integrated with the current state-of-the-art based on family history ascertainment, without substantially increasing the number of individuals identified as high risk.

### Limitations of the study

This study has several limitations. First, the study population is confined to adults residing in Nevada, thus, it may not completely represent the broader US population, let alone global populations. Second, we relied on three different sources in the EHR to obtain family history information, but it remains difficult to disentangle reasons why family history may be missing or when it is positive whether it is based on NCCN guidelines. Third, our analysis did not capture all individuals with monogenic risk. Specifically, our variant annotation and classification process did not evaluate many rare missense variants, particularly those in *PALB2*, *ATM* and *CHEK2*. We also did not evaluate variants in additional genes, such as *BARD1*, *RAD51C*, or *RAD51D*, which could increase the risk of breast cancer. These genes were not included despite being added in 2022 to the BOADICEA algorithm^41^ because (i) there was no statistical significant association between rare coding or rare loss-of-function variants in these genes and breast cancer when looking in the general population^13^, and (ii) other studies such as the WISDOM have yet to include these genes in their protocol^42^. Lastly, there may be biases in the accuracy of the polygenic risk score calculations based on genetic similarity. Making sure that polygenic risk scores have clinical utility for each individual and group of individuals is an important consideration to ensure the clinical utility of this tool for population screening ^40^. These limitations should be considered when interpreting the results of this study and planning future research to implement new strategies to identify women at a higher risk of breast cancer.

## Materials and Methods

### Subjects

This study was based on the Healthy Nevada Project. The Healthy Nevada Project study was reviewed and approved by the University of Nevada, Reno Institutional Review Board (IRB, project 956068-12), and all participants provided informed consent. The initial dataset comprised 39,546 individuals. For this study, we only included participants who were inferred to be of female sex based on the genetic data and had longitudinal data length >0 in the electronic health records at Renown Health.

### Clinical phenotypes from EHR

Phenotypes were processed from Epic/Clarity Electronic Health Records (EHR) data. Microsoft SQL Server was used as a backend database for record storage. SAS 9.4 M5 with SAS/ACCESS to SQL Server was used to perform ETL on these data and preparation for analysis, also in SAS using Base SAS and SAS/STAT. For this study, we focused on breast cancer and did not look at ovarian cancer despite the known impact of pathogenic variants in *BRCA1* and *BRCA2* on both breast and ovarian cancers. The mean number of ICD10-CM codes recorded per participant was 90 (median: 67).

ICD10-CM codes C50, D05, Z85.3 were used to identify women diagnosed with breast cancer. We did not take into account secondary neoplasm of the breast. For the phenotypes used as controls, we used the following ICD10-CM codes: codes starting with E10 for Type 1 diabetes, codes starting with E11 for Type 2 diabetes, codes starting with I10 for hypertension, and codes starting with M06 for rheumatoid arthritis. For family history of breast cancer, we used ICD10-CM code Z80.3. For family history of other conditions, we first excluded all Z80.3 codes, and then included all codes starting with Z80, Z81, Z82, Z83 or Z84.

### Family history information

Three sources of family history records for breast cancer were identified in the EHR: diagnosis codes, a family history table (FAMILY_HX), and a table containing responses to the seven-question family history screening (DM_BREAST_HEALTH_HX). Using data from August 2021, we evaluated the agreement between these three sources for identifying patients with a family history of breast cancer (**Figure S2A**). The diagnosis codes source was the most comprehensive. The majority of patients with indication of family history of breast cancer in the diagnosis codes did not have an indication in FAMILY_HX or DM_BREAST_HEALTH_HX tables, but 93.9% of those with indication of family history of breast cancer in FAMILY_HX and 100% of those with indication in DM_BREAST_HEALTH_HX also had a diagnosis code (**Figure S2A**).

We further evaluated whether using the FAMILY_HX table in addition to diagnosis codes would meaningfully change our conclusions about the effectiveness of family history for evaluating risk of breast cancer. To do this, we examined the temporal association of family history documentation with a diagnosis of breast cancer in two ways: determining dates of family history of breast cancer using (i) diagnosis codes only or using (ii) both diagnosis codes and FAMILY_HX. The difference in temporal association when FAMILY_HX is included or excluded appears to be negligible (**Figure S2B**). As a further test, we examined the temporal association between entries in FAMILY_HX and breast cancer without including diagnosis codes, to see if entries in FAMILY_HX also tended to appear after breast cancer diagnosis had already been diagnosed. We found that among 77 patients with both breast cancer and a breast cancer entry in the FAMILY_HX table, only 23.4% (18 of 77) had their 1st breast cancer entry in FAMILY_HX prior to their diagnosis of breast cancer (**Figure S2C**). 48% (28 of 59) of patients with their 1st breast cancer entry in the FAMILY_HX table after their breast cancer diagnosis had a family entry for another condition prior to the breast cancer diagnosis. These results validated the results reported in the main text (using FHx diagnosis codes only) showing that family history of breast cancer often fails to be documented prior to breast cancer, even if family history was previously assessed in some manner.

Based on these analyses, we concluded that diagnosis codes were sufficient for our purposes in evaluating the effectiveness of family history for breast cancer risk screening. Thus, for ease of reproducibility, we only utilize diagnosis codes in our main analysis.

### Exome+® sequencing

The HNP samples were sequenced at Helix using the Exome+® assay. Data was processed using a custom version of Sentieon and aligned to GRCh38, with variant calling and phasing algorithms following GATK best practices. Imputation of common variants in the HNP data was performed by pre-phasing samples and then imputing. Pre-phasing was performed using reference databases, which include the 1000 Genomes Phase 2 data. This was followed by genotype imputation for all 1000 Genomes Phase 3 sites that have genotype quality values <20.

### Variant annotation and classification

Variant annotation was performed with Ensembl Variant Effect Predictor-99^43^. The MANE transcripts were used to determine variant consequence^44^. Genotype processing was performed in Hail version 0.2.115-10932c754edb (https://github.com/hail-is/hail/commit/10932c754edb). Our strategy to identify variants increasing the risk of breast cancer (sometimes labeled as pathogenic or likely pathogenic) was based on two philosophical directions: (i) our study is focused on population-based screening and that many of the ACMG criteria that would apply for a diagnostic test do not apply or cannot be calculated in the context of screening a healthy individual^45^, and (ii) this is a research study and an automated variant annotation is more reproducible than a manual interpretation while still providing very similar results.

We focused on identifying variants in five genes: *BRCA1*, *BRCA2*, *PALB2*, *ATM* and *CHEK2*. While some studies only look at the common del1100C variant in *CHEK2*, we decided to analyze all *CHEK2* predicted loss-of-function variants. The following steps were done to annotate variants and identify loss-of-function variants:

1. Preparation of the genetic file: restrict Hail matrix table to specific genomic intervals for the five genes.
2. Annotate with VEP, with Clinvar (file clinvar_20220723.vcf.gz downloaded from https://ftp.ncbi.nlm.nih.gov/pub/clinvar/vcf_GRCh38/ was used) and with gnomAD_v3 (https://gnomad.broadinstitute.org/downloads#v3-variants). There were 13,853 variants in the 5 genes before any filtering.
3. Filter out variants flagged as ‘Filtered variants’ by gnomAD because they did not pass their quality control process. 13,673 variants remained.
4. We then did the annotation using the following logic:

a. IF the variant was rs555607708 CHEK2 del1100C variant THEN label as ‘P/LP’.
b. ELSE IF the variant was reviewed by the Clingen expert panel (CLNREVSTAT field in Clinvar table) THEN keep the interpretation from the Clingen expert panel.
c. ELSE IF the variant had criteria provided by multiple submitters with no conflicts (CLNREVSTAT field in Clinvar table) and was ‘Benign’ or ‘Likely Benign’ (CLNSIG field) THEN label as ‘B/LB’.
d. ELSE IF the variant had criteria provided by multiple submitters with no conflicts (CLNREVSTAT field in Clinvar table) and was ‘Uncertain significance’ (CLNSIG field) THEN label as ‘VUS.
e. ELSE IF the variant was called as LoF with High Confidence by LOFTEE^46^, THEN label ‘P/LP’. Briefly, LOFTEE flags LoF variants as low confidence (LC) if the LoF version is the ancestral state, if they are a stop gain or frameshift near the end of the gene or are in an exon with non canonical splice sites around it, or if they are a splice variant that is not predicted to affect the splicing of a coding exon.
f. ELSE label variant as ‘not pathogenic’.
5. A total of 184 variants were called ‘P/LP’ this way.
6. Additional quality control was done for each of these variants including review of the DP, AD, GQ fields, the AF in our cohort and in other databases and visualization of the BAM file with IGV for small insertions or deletions.
7. Two variants in the same gene and same individual were removed as likely false positives after reviewing the BAM file. A total of 182 variants listed in **Table S1** were annotated as pathogenic variants for this study.

### CNV annotations

The Helix Exome+® assay includes a copy-number variant (CNV) caller, allowing us to incorporate rare CNVs at exon-level resolution. Briefly, CNVs with the PASS QC filter were annotated with overlapping MANE transcripts. Only large deletions were considered to be pathogenic for this study. The list of CNVs identified as pathogenic is in **Table S2**.

### Polygenic risk score calculation

The PRS model selected is the 313 SNPs PRS published in 2019^14^. It is also available in the PGS catalog^36^ (https://www.pgscatalog.org/score/PGS000004/). We first converted the coordinates and effect size of each alternate allele from human reference genome GRCh37 to the more recent GRCh38 (**Table S5**). We used 300 SNPs out of the 313 to ensure we had strong overall callability for each SNP used and confidence the alternate (and effect) allele were correct. Allele frequencies of alternative alleles matched closely with published allele frequencies for these variants. **(Figure S4B**).

We then calculated the score in each of the 25,591 women included in the study. The distribution of genetic similarity was the following: N Africa = 499, N Americas = 3,728, N East Asia = 832, N Europe = 19,484, N Other = 929 and N South Asia = 119. Briefly, a genotype dosage was calculated for each variant in the score for each individual. The dosage was based on the genotype probability field resulting from the imputation pipeline. When an individual had no GP or no GT (genotype) for a specific variant, the dosage was based on the Allele Frequency of this variant in gnomAD v3 for the population closest to the genetic similarity of the participant. We then split the cohort into 6 cohorts based on genetic similarity and ranked individuals based on their PRS value, and assigned a percentile based on the ranking within the participant’s genetic similarity distribution.

Lastly, we regrouped all six genetic similarity groups into one cohort for later analyses that were based on percentiles.

### Cancer risk thresholds

Different definitions and thresholds exist to stratify women considered at high risk of breast cancer.

- The National Comprehensive Cancer Network (NCCN) guidelines consider ‘Increased risk’ an asymptomatic women with a residual lifetime risk ≥20% as defined by models that are largely based on family history^18^.
- The American Cancer Society considers women at high risk those that have a lifetime risk of breast cancer of about 20% to 25% or greater, according to risk assessment tools that are based mainly on family history^17^.
- the UK NICE (National Institutes of Clinical and Healthcare Excellence) guidelines use a threshold of >30% at age 80 for ‘high risk’, and between 17 and 30% at age 80 for ‘moderate risk’^16^.

Here, we followed the NCCN definition and considered a woman to be at increased risk, or high risk if they had an accumulated risk of being diagnosed with breast cancer ≥20% by age 70.

### Survival analysis, Hazard ratios and statistical tests

The specific tests used are described in detail in the main text or in the figure legends.

Kaplan Meier survival curves were done using the KaplanMeierFitter function from the Lifelines python library.

Statistical differences between survival curves were assessed using a logrank_test function from the lifelines.statistics python library.

Values at a given age (e.g. 70 years old) were calculated using the ‘predict’ function.

Hazard ratios were calculated using the CoxPHFitter function from the lifelines python library. Plots were made using pyplot from the matplotlib python library.

## Data Availability

The Healthy Nevada Project data are available to qualified researchers upon request and with permission of the Institute for Health Innovation (IHI) and Helix. Researchers who would like to obtain the raw genotype data related to this study will be presented with a data user agreement, which requires that no participants will be reidentified and no data will be shared between individuals or uploaded onto public domains. The IHI encourages and collaborates with scientific researchers on an individual basis. Examples of restrictions that will be considered in requests to data access include but are not limited to (1) whether the request comes from an academic institution in good standing and will collaborate with our team to protect the privacy of the participants and the security of the data requested, (2) type and amount of data requested, (3) feasibility of the research suggested, and (4) amount of resource allocation for the IHI and Renown Hospital required to support the collaboration. Any correspondence and data availability requests related to Healthy Nevada Project should be addressed to J.J.G. or Craig Kugler.

## Acknowledgments

We thank the participants in the Healthy Nevada Project. We thank all of the staff organizing and running the Healthy Nevada Project. Funding was provided to Desert Research Institute (DRI) by the Nevada Governor’s Office of Economic Development. Funding was provided to the Renown Institute for Health Innovation by Renown Health and the Renown Health Foundation.We also thank the laboratory and bioinformatics teams at Helix, as well as the clinical / medical affairs team for helpful discussions. We thank Catherine Clinton and Lexa Mackie for assistance with the IRB.

## Author Information

Conceptualization: A.B., J.J.G.; Data curation: K.M.S.B., R.J., I.T., N.T., R.J.; Formal analysis: A.B., D.K.; Resources: G.E., I.N., S.D., H.R., A.A, W.J.M., N.L.W.; Supervision: E.T.C., J.J.G.; Writing-original draft: A.B.; Writing-review and editing: A.B., D.K., K.M.S.B., G.E., J.M.S.B., E.O., M.J.F., C.H., E.T.C., J.J.G.

## Ethics Declaration

The Healthy Nevada Project study was reviewed and approved by the University of Nevada, Reno Institutional Review Board (project 956068-12). All participants gave their informed consent before participation. All data used for research were de-identified.

## Conflict of Interest

A.B., K.M.S.B., I.T., N.T., R.J., N.L.W., M.J.F., C.H. and E.T.C. are employees of Helix. A patent application has been filed by Helix for the ‘Dynamic risk management for breast cancer based on multi-factor genetic testing’ with A.B. and J.J.G. as inventors, and its current status is unpublished (application number 63/467,250).

## Supplementary Information

There are 5 supplementary figures and 9 supplementary tables.

**Figure S1:**
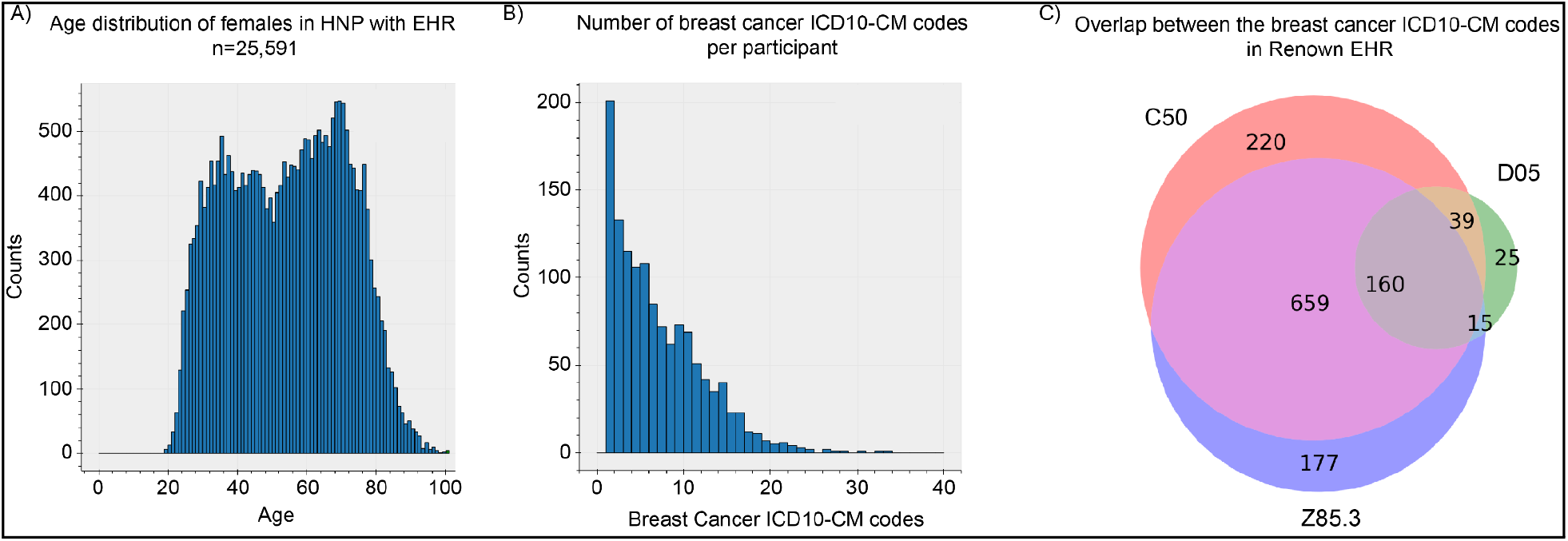
Clinical characteristics of the research participants included in this study. **(A)** Distribution of age in 2023 in the HNP female participants. **(B)** Number of entries of ICD10-CM codes indicating a breast cancer diagnosis (ICD10-CM codes starting with C50, D05, or Z85.3) per participant among women with at least 1 of these codes. **(C)** Number of women with at least one of each ICD10-CM code.

**Figure S2:**
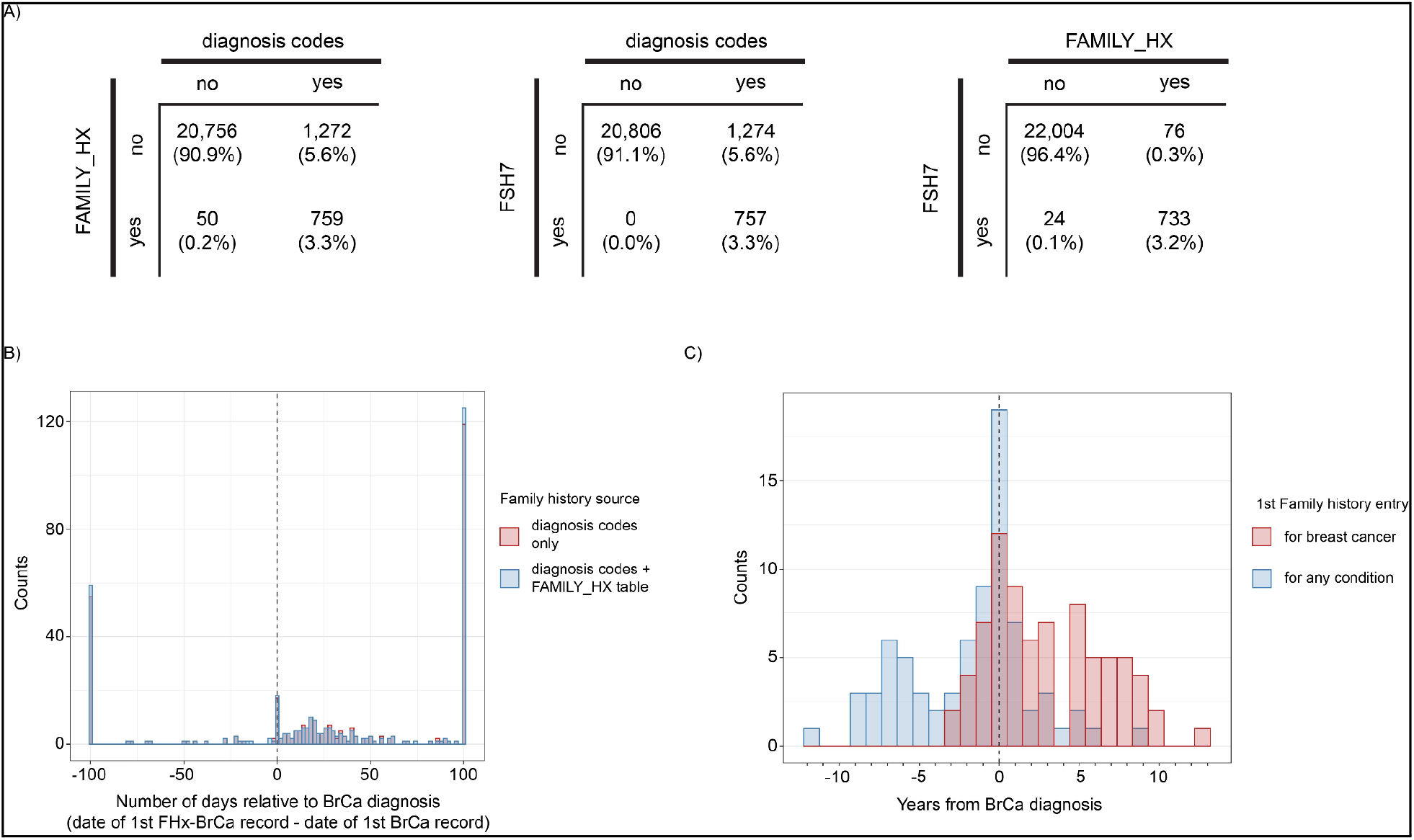
Analysis of sources of breast cancer family history records in the EHR. Analysis done using data from 22,837 HNP participants from August 2021. **(A)** Confusion matrices indicating correspondence between data sources documenting a family history of breast cancer (“no” indicating no documentation, “yes” indicating a positive documentation). FAMILY_HX is the family history table. FSH7 represents the table containing responses to the seven-question family history screening (table also called: DM_BREAST_HEALTH_HX). **(B)** Temporal distribution of initial documentation of family history for breast cancer relative to date of initial breast cancer diagnosis – a comparison of when only diagnosis codes are used versus when entries from the FAMILY_HX table are also incorporated. N=334 for diagnosis codes only and N=344 for diagnosis codes + FAMILY_HX table. **(C)** Temporal distribution of initial FAMILY_HX table entries for breast cancer versus the temporal distribution of initial FAMILY_HX table entries for any condition, relative to date of breast cancer diagnosis. N=77.

**Figure S3:**
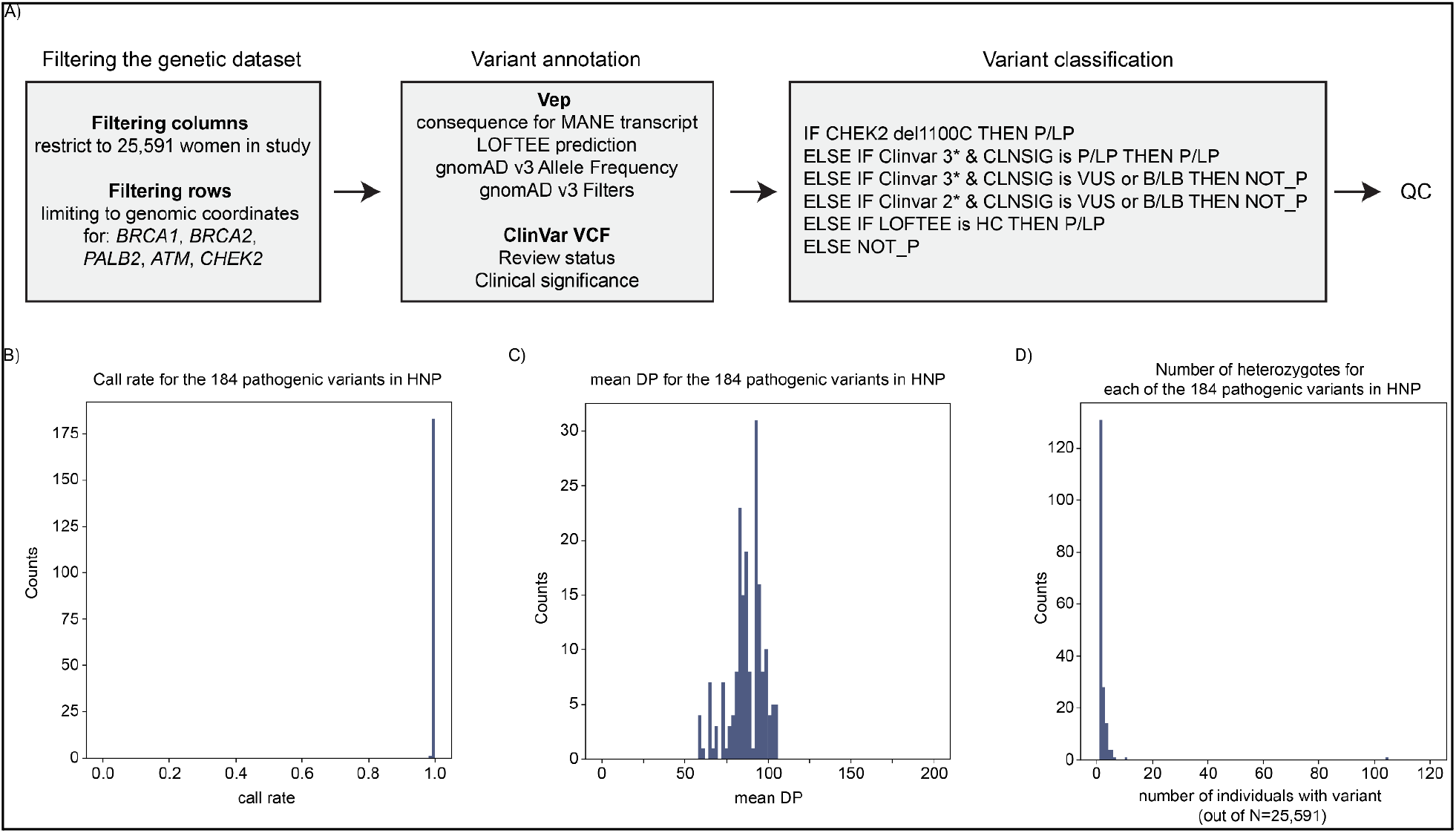
Process for variant annotation, classification and variant call quality control. **(A)** Schematic of the workflow for variant annotation and classification. References to some of the tools used are provided in the Material and Methods section. Clinvar 3* represents a Clinvar review status of ‘Reviewed by Clingen expert panel’. Clinvar 2* represents a Clinvar review status of ‘Multiple submitters, no conflicts’. **(B)** Call rate in the study cohort for the 184 variants classified as ‘pathogenic’. **(C)** Mean DP (read depths) for the 184 variants classified as ‘pathogenic’. **(D)** Number of women heterozygotes for each of the 184 variants classified as ‘pathogenic’. The one variant with more than 100 individuals with the variant is *CHEK2* del1100C.

**Figure S4:**
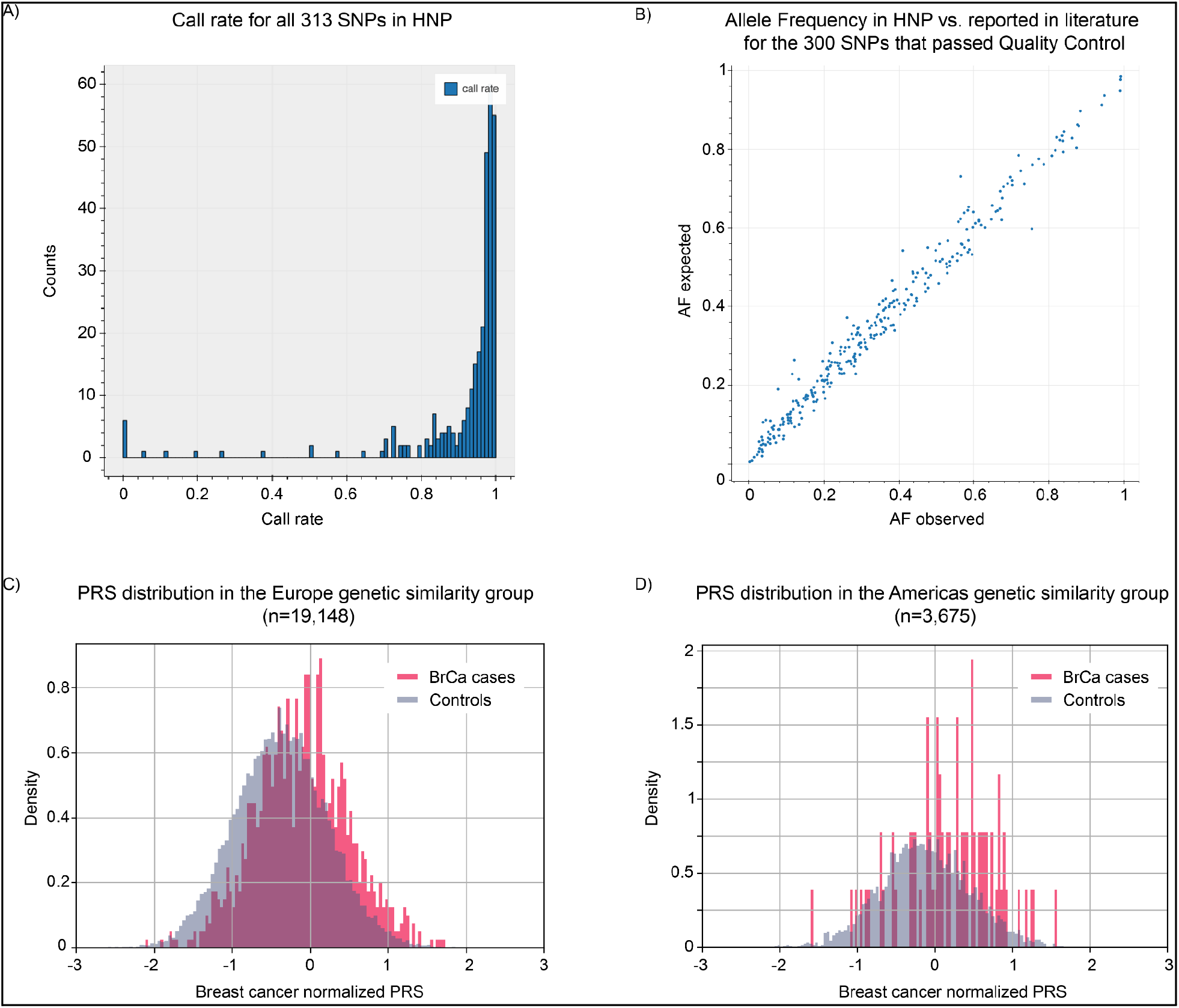
Implementation of the breast cancer polygenic risk score. (A) Call rate in HNP for the 313 SNPs included in the PRS model used in this study^14^. **(B)** Scatter plot showing the expected Allele Frequency (AF) compared to the observed AF in the Healthy Nevada Project participants. The expected AF comes from the original paper that published this score^14^. Each dot is one of the 300 SNPs that passed our internal quality control. **(C and D)** Distribution of the normalized PRS values in breast cancer cases (pink) and in controls (blue). Panel C represents individuals with Europe genetic similarity. Panel D represents individuals with Americas genetic similarity.

**Figure S5:**
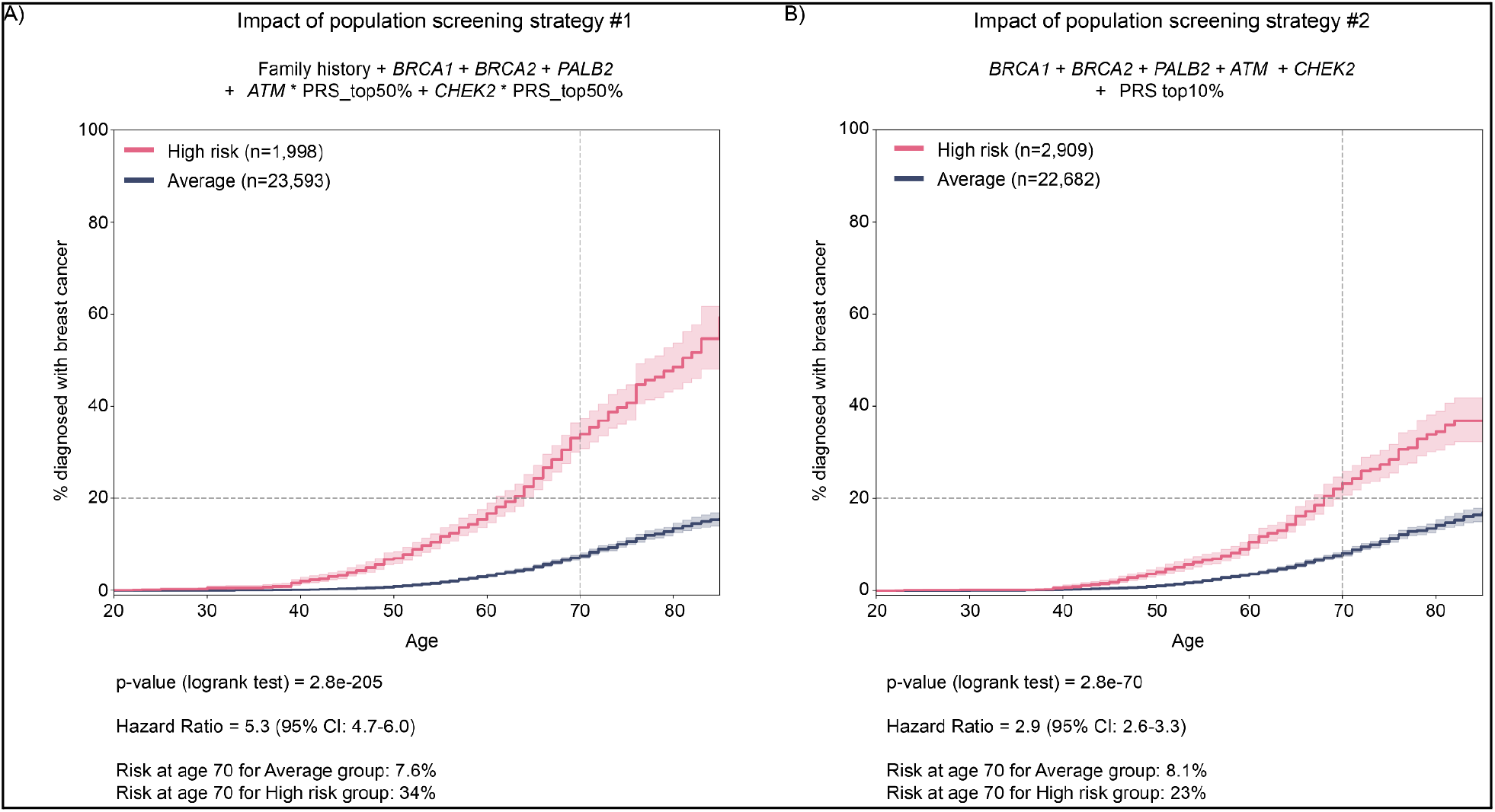
Impact of two potential strategies to identify women at high risk of breast cancer in the population. **(A)** Kaplan Meier curves showing the % of women with a breast cancer diagnosis by age based on whether they were identified as ‘high risk’ (pink curve) or at average risk (blue curve) with strategy #1. **(B)** Kaplan Meier curves showing the % of women with a breast cancer diagnosis by age based on whether they were identified as ‘high risk’ (pink curve) or at average risk (blue curve) with strategy #2.

**Table S1:** List of all variants classified as pathogenic.

**Table S2:** List of all copy number variants classified as pathogenic.

**Table S3:** Number of individuals with a pathogenic variant per gene.

| Gene | N carriers total | By consequence | With a Breast Cancer diagnosis |
| --- | --- | --- | --- |
| *BRCA1* | 66 | Stop gained: 14<br>Frameshift: 32<br>Splice acceptor or donor: 3<br>Missense, splice region, start lost: 12<br>Deletion (copy number of 1) of 1 exon or more: 5 | 25 (38%) |
| *BRCA2* | 94* | Stop gained: 13<br>Frameshift: 73<br>Splice acceptor or donor: 3<br>Missense, splice region, start lost: 2<br>Deletion (copy number of 1) of 1 exon or more: 3 | 27 (29%)* |
| *PALB2* | 39 | Stop gained: 15<br>Frameshift: 17<br>Splice acceptor or donor: 1<br>Missense, splice region, start lost: 0<br>Deletion (copy number of 1) of 1 exon or more: 6 | 10 (26%) |
| *ATM* | 73 | Stop gained: 14<br>Frameshift: 32<br>Splice acceptor or donor: 15<br>Missense, splice region, start lost: 4<br>Deletion (copy number of 1) of 1 exon or more: 8 | 15 (21%) |
| *CHEK2* | 139 | Stop gained: 1<br>Frameshift: 124 (including 104 of *CHEK2* del1100C variant).<br>Splice acceptor or donor: 5<br>Missense, splice region, start lost: 0<br>Deletion (copy number of 1) of 1 exon or more: 9 | 18 (13%) |
| none | 25,181 |  | 1,200 (4.8%) |

\*One participant carried both a *BRCA1* and a *BRCA2* pathogenic variant. For the phenotype analysis, this participant was considered to have a *BRCA1* pathogenic variant.

**Table S4:** Clinical impact of pathogenic variants on breast cancer diagnosis.

| Gene | HR | CI | p-value | Probability of diagnosis at age 70 | Age of earliest diagnosis |
| --- | --- | --- | --- | --- | --- |
| BRCA1 | 16.2 | 10.9-24.1 | 2.6e-76 | 76% | 29 |
| BRCA2 | 8.5 | 5.8-12.5 | 1.3e-39 | 55% | 22 |
| PALB2 | 6.3 | 3.4-11.7 | 3.2e-11 | 36% | 23 |
| ATM | 4.3 | 2.6-7.2 | 1.0e-09 | 37% | 39 |
| CHEK2 | 2.6 | 1.7-4.2 | 2.4e-05 | 19% | 47 |
| None | 1 | NA | NA | 9.3% | 20 |

**Table S5:** SNPs and coordinates in GRCh38 for breast cancer polygenic risk score.

**Table S6:** Median breast cancer polygenic risk score for different populations grouped by genetic ancestry.

| Genetic similarity group | Cutoff for lower 10% | Median PRS | Cutoff for top 10% |
| --- | --- | --- | --- |
| Africa | -0.70 | 0.09 | 0.84 |
| Americas | -0.84 | -0.13 | 0.64 |
| East Asia | -0.45 | 0.21 | 0.90 |
| Europe | -1.11 | -0.36 | 0.40 |
| Other | -0.89 | -0.08 | 0.68 |
| South Asia | -1.24 | -0.28 | 0.29 |

**Table S7:** Distribution and performance of the score in HNP.

| PRS decile | N total | N cases | N controls | Fraction of cases | N with positive family history (recorded at any time) | Fraction with positive family history |
| --- | --- | --- | --- | --- | --- | --- |
| 0 | 2,529 | 54 | 2,475 | 0.022 | 109 | 0.043 |
| 10 | 2,518 | 64 | 2,454 | 0.026 | 133 | 0.053 |
| 20 | 2,522 | 80 | 2,442 | 0.033 | 144 | 0.057 |
| 30 | 2,515 | 103 | 2,412 | 0.043 | 153 | 0.061 |
| 40 | 2,503 | 104 | 2,399 | 0.043 | 161 | 0.064 |
| 50 | 2,524 | 107 | 2,417 | 0.044 | 190 | 0.075 |
| 60 | 2,527 | 116 | 2,411 | 0.048 | 191 | 0.076 |
| 70 | 2,530 | 166 | 2,364 | 0.07 | 193 | 0.076 |
| 80 | 2,514 | 169 | 2,345 | 0.072 | 201 | 0.08 |
| 90 | 2,499 | 237 | 2,262 | 0.11 | 211 | 0.084 |

**Table S8:**
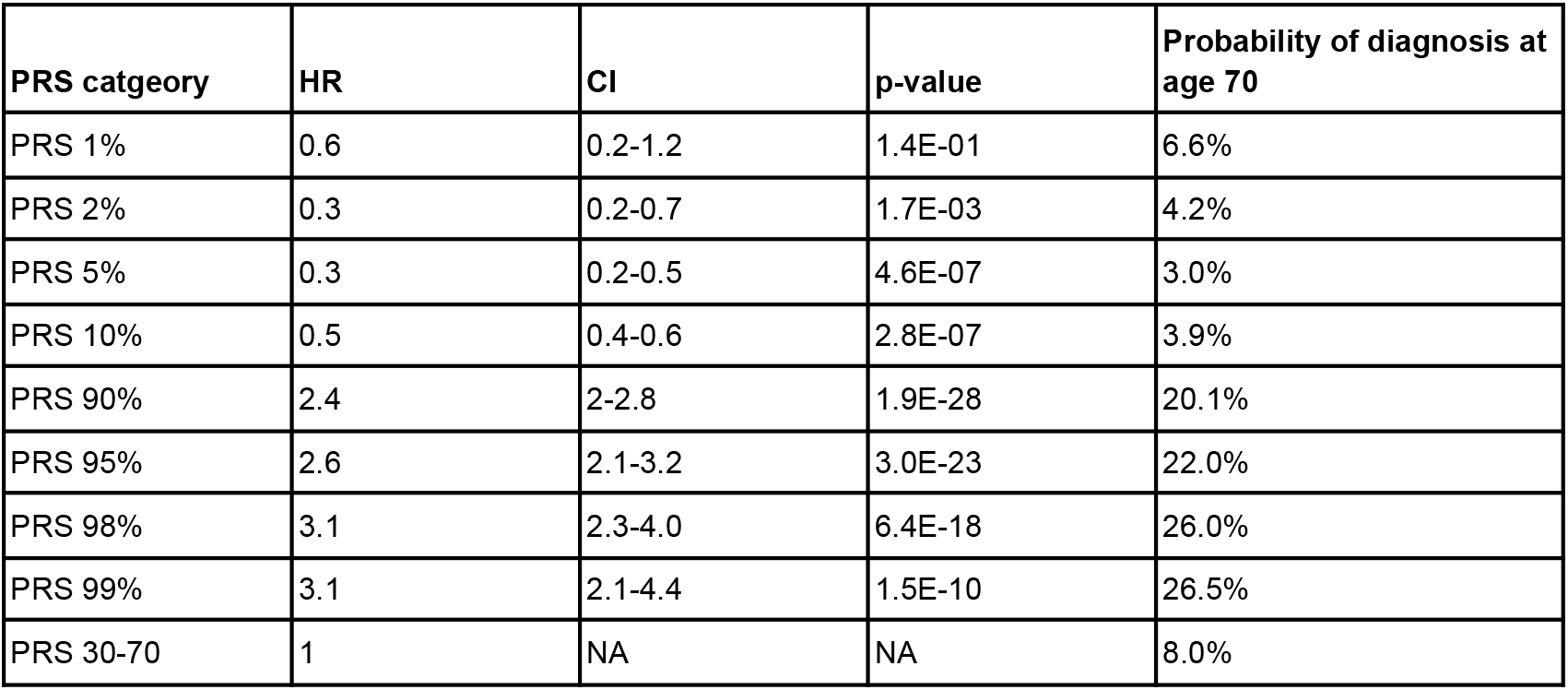
Clinical impact of the polygenic risk score on breast cancer diagnosis.

| PRS catgeory | HR | CI | p-value | Probability of diagnosis at age 70 |
| --- | --- | --- | --- | --- |
| PRS 1% | 0.6 | 0.2-1.2 | 1.4E-01 | 6.6% |
| PRS 2% | 0.3 | 0.2-0.7 | 1.7E-03 | 4.2% |
| PRS 5% | 0.3 | 0.2-0.5 | 4.6E-07 | 3.0% |
| PRS 10% | 0.5 | 0.4-0.6 | 2.8E-07 | 3.9% |
| PRS 90% | 2.4 | 2-2.8 | 1.9E-28 | 20.1% |
| PRS 95% | 2.6 | 2.1-3.2 | 3.0E-23 | 22.0% |
| PRS 98% | 3.1 | 2.3-4.0 | 6.4E-18 | 26.0% |
| PRS 99% | 3.1 | 2.1-4.4 | 1.5E-10 | 26.5% |
| PRS 30-70 | 1 | NA | NA | 8.0% |

**Table S9:**
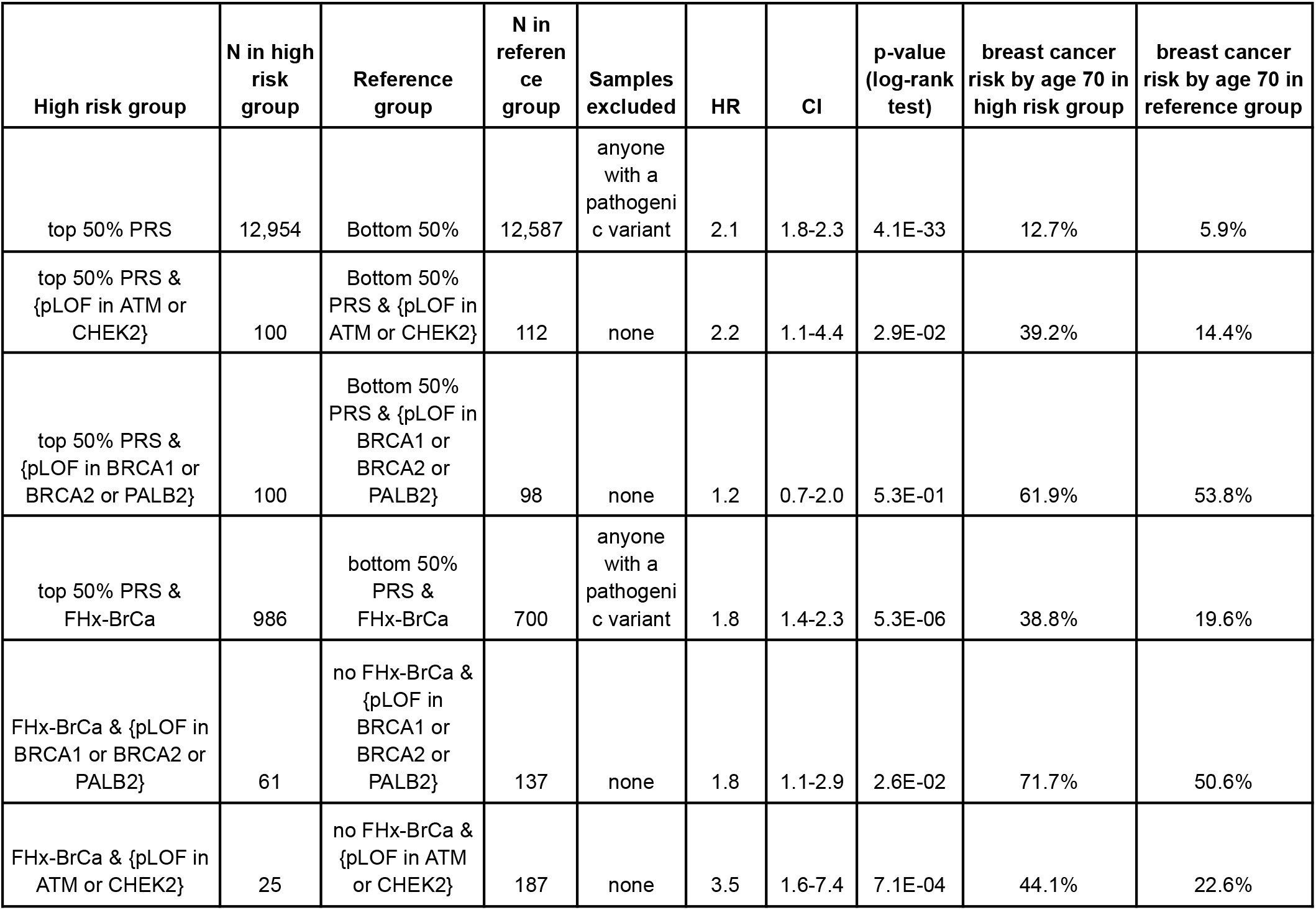
Impact of combining two non-modifiable risk factors on breast cancer diagnosis.

| High risk group | N in high risk group | Reference group | N in reference group | Samples excluded | HR | CI | p-value (log-rank test) | breast cancer risk by age 70 in high risk group | breast cancer risk by age 70 in reference group |
| --- | --- | --- | --- | --- | --- | --- | --- | --- | --- |
| top 50% PRS | 12,954 | Bottom 50% | 12,587 | anyone with a pathogenic variant | 2.1 | 1.8-2.3 | 4.1E-33 | 12.7% | 5.9% |
| top 50% PRS & {pLOF in ATM or CHEK2} | 100 | Bottom 50% PRS & {pLOF in ATM or CHEK2} | 112 | none | 2.2 | 1.1-4.4 | 2.9E-02 | 39.2% | 14.4% |
| top 50% PRS & {pLOF in BRCA1 or BRCA2 or PALB2} | 100 | Bottom 50% PRS & {pLOF in BRCA1 or BRCA2 or PALB2} | 98 | none | 1.2 | 0.7-2.0 | 5.3E-01 | 61.9% | 53.8% |
| top 50% PRS & FHx-BrCa | 986 | bottom 50% PRS & FHx-BrCa | 700 | anyone with a pathogenic variant | 1.8 | 1.4-2.3 | 5.3E-06 | 38.8% | 19.6% |
| FHx-BrCa & {pLOF in BRCA1 or BRCA2 or PALB2} | 61 | no FHx-BrCa & {pLOF in BRCA1 or BRCA2 or PALB2} | 137 | none | 1.8 | 1.1-2.9 | 2.6E-02 | 71.7% | 50.6% |
| FHx-BrCa & {pLOF in ATM or CHEK2} | 25 | no FHx-BrCa & {pLOF in ATM or CHEK2} | 187 | none | 3.5 | 1.6-7.4 | 7.1E-04 | 44.1% | 22.6% |

